# Reagent-free Hyperspectral Diagnosis of SARS-CoV-2 Infection in saliva samples

**DOI:** 10.1101/2024.06.03.24308300

**Authors:** Brandon Saint-John, Alejandro Wolf-Yadlin, Daniel E. Jacobsen, Jamie L. Inman, Serge Gart, Matt Keener, Cynthia McMurray, Antoine M. Snijders, Harshini Mukundan, Jessica Z. Kubicek-Sutherland, James B. Brown

**Author notes:** Corresponding authors: -Harshini Mukundan, Lawrence Berkeley National Laboratory 1 Cyclotron Road, Berkeley, CA 94720, -Jessica Z. Kubicek-Sutherland Los Alamos National Laboratory PO Box 1663, MS J567, Los Alamos, NM 87545, -Antoine M. Snijders, Lawrence Berkeley National Laboratory 1 Cyclotron Road, Berkeley, CA 94720, -Matt Keener, Pattern Computer, Inc. 4020 148^th^ Ave NE, Bldg N Ste B Redmond, WA, 98052. These authors contributed equally (Brandon Saint-John, Alejandro Wolf-Yadlin, Daniel E. Jacobsen).

## Abstract

**Background:** Rapid, reagent-free pathogen-agnostic diagnostics that can be performed at the point of need are vital for preparedness against future outbreaks. Yet, many current strategies (polymerase chain reaction, lateral flow immunoassays) are pathogen-specific and require reagents; whereas others such as sequencing-based methods; while agnostic, are not (as yet) conducive for use at the point of need. Herein, we present hyperspectral sensing as an opportunity to overcome these barriers, realizing truly agnostic reagent-free diagnostics. This approach can identify both pathogen and host signatures, without complex logistical considerations, in complex clinical samples. The spectral signature of biomolecules across multiple wavelength regimes provides rich biochemical information, which, coupled with machine learning, can facilitate expedited diagnosis of disease states, the feasibility of which is demonstrated here.

**Innovation:** First, we present ProSpectral™ V1, a novel, miniaturized (∼8 lbs) hyperspectral platform with ultra-high (2-5 nm full-width, half-max, i.e., FWHM) spectral resolution that incorporates two mini-spectrometers (visual and near-infrared). This engineering innovation has enabled reagent-free biosensing for the first time. To enable expedient outcomes, we developed state-of-the-art machine learning algorithms for near real-time analysis of multi-wavelength spectral signatures in complex samples. Taken together, these innovations enable near-field ready, reagent-free, expedient agnostic diagnostics in complex clinical samples. Herein, we demonstrate the feasibility of this synergy of ProSpectral™ V1 with machine learning to accurately identify

SARS-CoV-2 infection status in double-blinded saliva samples in real-time (3 seconds/measurement). The infection status of the samples was validated with the CDC-approved polymerase-chain reaction (PCR). We report accuracies comparable to first-in-class PCR tests. Further, we provide preliminary support that this signal is specific to SARS-CoV-2, and not associated with other respiratory conditions.

**Interpretation:** Preparedness against unanticipated pathogens and democratization of diagnostics requires moving away from technologies that demand specific reagents; and relying on intrinsic biochemical properties that can, theoretically, inform on *all* pathologies. Integration of hyperspectral sensors and in-line machine learning analytics, as reported here, shows the feasibility of such diagnostics. If realized to full potential, the ProSpectral™ V1 platform can enable agnostic diagnostics, thereby improving situational awareness and decision-making at the point of need; especially in resource-limited settings – enabling the distribution of newly developed tests for emerging pathogens with only a simple software update.

**Funding:** The U.S. Department of Energy, the Defense Threat Reduction Agency, Lawrence Berkeley National Laboratory, Los Alamos National Laboratory, and Pattern Computer Inc.

**Research in context:** *Evidence before this study:* Our inability to quickly and effectively deploy and use reliable diagnostics at the point of need is a major limitation in our arsenal against infectious diseases. We searched PubMed and Google Scholar for articles published before May 2024 in English applying hyperspectral sensing technologies of pathogen detection with terms, “hyperspectral,” “pathogens”, and “COVID-19”. Various factors such as speed, sensitivity, availability of reagents, deployability, requirements (expertise, resources), and others determine our choice of diagnostic. Today, diagnosis of infection remains largely pathogen-specific, requiring ligands specific to the target of interest. Indeed, Polymerase Chain Reaction (PCR)-based methods, the gold-standard technology to diagnose COVID-19, are pathogen-specific and have to be re-evaluated with the emergence of new variants. Lateral flow immunoassays, while readily deployable, are associated with lower sensitivity and specificity, and require the development of ligands, which can be time-consuming when addressing unanticipated or new threats. Select pathogen-agnostic methods such as sequencing are evolving and becoming more feasible, but still require sample processing, reagents, cold-chain, and expert handlers - and hence are not (as yet) available for routine point-of-care use. In contrast, the characterization of biochemical signatures across multiple spectral regimes (hyperspectral) can facilitate reagent-free agnostic diagnostics. Yet, many spectroscopic methods are either limited to narrow wavelength ranges; or are too large for use in the point-of-care setting; and may require complex and time-consuming analytics.

*Added value of this study:* This manuscript presents a paradigm-shifting miniaturized hyperspectral sensor with embedded machine learning-enabled analytics that can overcome the above limitations, making reagent-free agnostic diagnostics achievable. To our knowledge, this establishes the fastest hyperspectral diagnostic platform (3 seconds/measurement), with no preprocessing and in a small form factor, and executable with liquid (clinical) samples, without ligands or reagents. Our data demonstrates that the sensitivity of this assay is comparable to gold-standard PCR-based assays; and that the signatures are specific to COVID-19 and not associated with influenza and other respiratory pathogens – establishing the truly agnostic nature of the platform. The sensor consists of two embedded spectrometers, covering spectral bandwidth 400-1700 nm, which covers spectral patterns associated with relevant biological moieties. With appropriate data processing, we demonstrate balanced accuracies between 0·97 and 1·0 under a 10-fold cross-validation (depending on the ML/AI algorithm used for prediction).

*Implications of all the available evidence:* With the optimization of algorithms and analytical methods and the development of appropriate spectral databases, the ProSpectral™ hyperspectral diagnostics platform can be a flexible tool for rapid, reagent-free pathogen-agnostic detection/diagnosis of disease at the point of need, which can be a disruptive force in our preparedness to counter emerging diseases and threats.

## Introduction

Rapid identification of infection at the point of need is critical to curbing downstream severity, incidence, mortality, and morbidity.^1,2^ In current practice, the choice of the biochemical target (e.g., nucleic acids vs. proteins), the mode of disease manifestation (e.g.; respiratory vs. gastrointestinal), and the availability of resources and reagents; all determine the type of diagnostic and its deployment across a population.^3^ During the COVID-19 pandemic, nucleic acid amplification methods such as PCR were identified as the gold standard for pathogen identification.^4^ Lateral flow immunoassays were deployed as home-based diagnostics for improved situational awareness, while PCR-based methods provided specific identification at the regional laboratory. While both methods are highly effective for diagnosing COVID and other diseases, they have significant drawbacks. With regards to the identification of new and emerging threats and future outbreaks, the most significant drawback of these methods is that they are pathogen-specific. While easily deployed and simple to use, lateral flow immunoassays require specific antibodies and/or ligands; and are also intrinsically associated with poor sensitivity compared to nucleic-acid amplification methods. The latter, such as PCR, are highly specific and sensitive. Yet, they require reagents, cold chain procedures, and, most commonly, a laboratory interface for execution. Finally, the specificity of a diagnostic, while desirable, requires constant iteration and re-design of primers and probes to address emerging variants, as was evidenced during the COVID-19 pandemic.^5,6^ Overall, conventional targeted diagnostics do not prepare us to rapidly address emerging and unanticipated outbreaks. Thus, there is an urgent need for pathogen-agnostic and readily deployable, easy-to-use, reagent-free platforms to effectively combat the next outbreak.

Beyond outbreaks, such technologies can also improve our ability to routinely identify and effectively treat infectious diseases in the clinical setting. Previous work from our laboratory and others has highlighted the risk of missed or ineffective diagnostics in routine medical practice and highlighted the need for effective methods for routine use in such situations.^4,7,8^ Sequencing-based methods have demonstrated the most promise of the pathogen-agnostic diagnostic modalities in development.^9^ Next-generation sequencing (NGS) technologies can (arguably) identify any pathogen in a clinical sample, including previously uncharacterized or emerging microbes. Studies from our team demonstrated the effectiveness of this method in accurately identifying respiratory pathogens associated with disease in patients.^4,10^ However, the dependency of the platform on reagents and expertise cannot be questioned, and there is a need for further innovation and automation to enable ready use at the point of need. Beyond sequencing, other biochemical sensor platforms are currently being studied.^11^ These include lab-on-a-chip and microfluidics-based methods, surface-enhanced Raman spectroscopy, and surface plasmon resonance-based methods; among others. ^12^ While many hold promise, the critical need for reagent-free pathogen-agnostic diagnostics at the point of need remains unresolved.

Herein, we present hyperspectral sensing as a potential solution to this significant challenge. Hyperspectral sensing is a non-destructive measurement technique that uses light to understand the composition of complex mixtures. In hyperspectral sensing, light at multiple sequential wavelengths is passed through a sample. The molecular bonds of the compounds in the sample will absorb light at specific wavelengths. A spectrometer measures the light that isn’t absorbed and compares it with the light input, and the difference at each wavelength forms spectra. All biological moieties emanate distinctive spectral signatures which can provide unique signature patterns that can theoretically allow for the identification of all biomarkers. Indeed, several studies have applied hyperspectral sensing to develop diagnostics. These have proven accurate in detecting biomarkers for Huntington’s disease, COVID-19, and more.^13–22^ However, most of these studies used traditional spectrometers, which are typically bulky and expensive. More recently, a new class of miniaturized spectrometers has enabled the adaptation of this technology for inexpensive point-of-care diagnostics.^23^ To take advantage of the size and speed of these miniature spectrometers, we also need to choose a sample collection approach that is rapid, low-cost, and readily available.

Saliva has been increasingly recognized as a promising, non-invasive sample type.^24^ Infectious disease, cancer - both localized and systemic, cardiovascular disease, diabetes, and more have demonstrated detectable biomarkers in saliva.^24–26^ We choose to use saliva as the diagnostic medium for detecting SARS-CoV-2 infection. Unlike previous studies that use saliva for diagnostics, our study did not employ any preprocessing steps to enhance data quality.

Thus, miniaturized hyperspectral sensing can provide a reagent-free platform for point-of-care pathogen-agnostic diagnostics via measurement of COVID-19 metabolites in saliva. While further work is required to demonstrate the full potential of this technology, the work shown here offers preliminary insights into the feasibility of realizing the same.

## Results

A miniaturized hyperspectral platform combining multiple spectrometers is ideal for efficiently capturing the desired spectra. Pattern’s ProSpectral^TM^ V1 platform combined custom software drivers and a data acquisition package deployed to a small, embedded computing system with two miniature spectrometers (Figure 1A). The resulting platform is capable of measuring biological moieties from 400-1700 nm, in continuum. At these wavelength ranges, all key biomolecular species – proteins, lipids, nucleic acids and carbohydrates – are captured effectively. To validate the real-world efficacy of this platform, we performed measurements in blinded saliva samples collected from a commercial source as described in the methods (Figure 1B). We first generated ten replicate spectral measurements from 470 saliva samples over the 400-1700 nm wavelength range. Our sample set includes male and female participants (Figure 2A, B), across multiple age groups. After quality processing (see methods), our dataset contained replicate hyperspectral measurements from 359 samples, split between 152 SARS-CoV-2-negative and 207 SARS-CoV-2-positive samples (Figure 2C).

**Figure 1.**
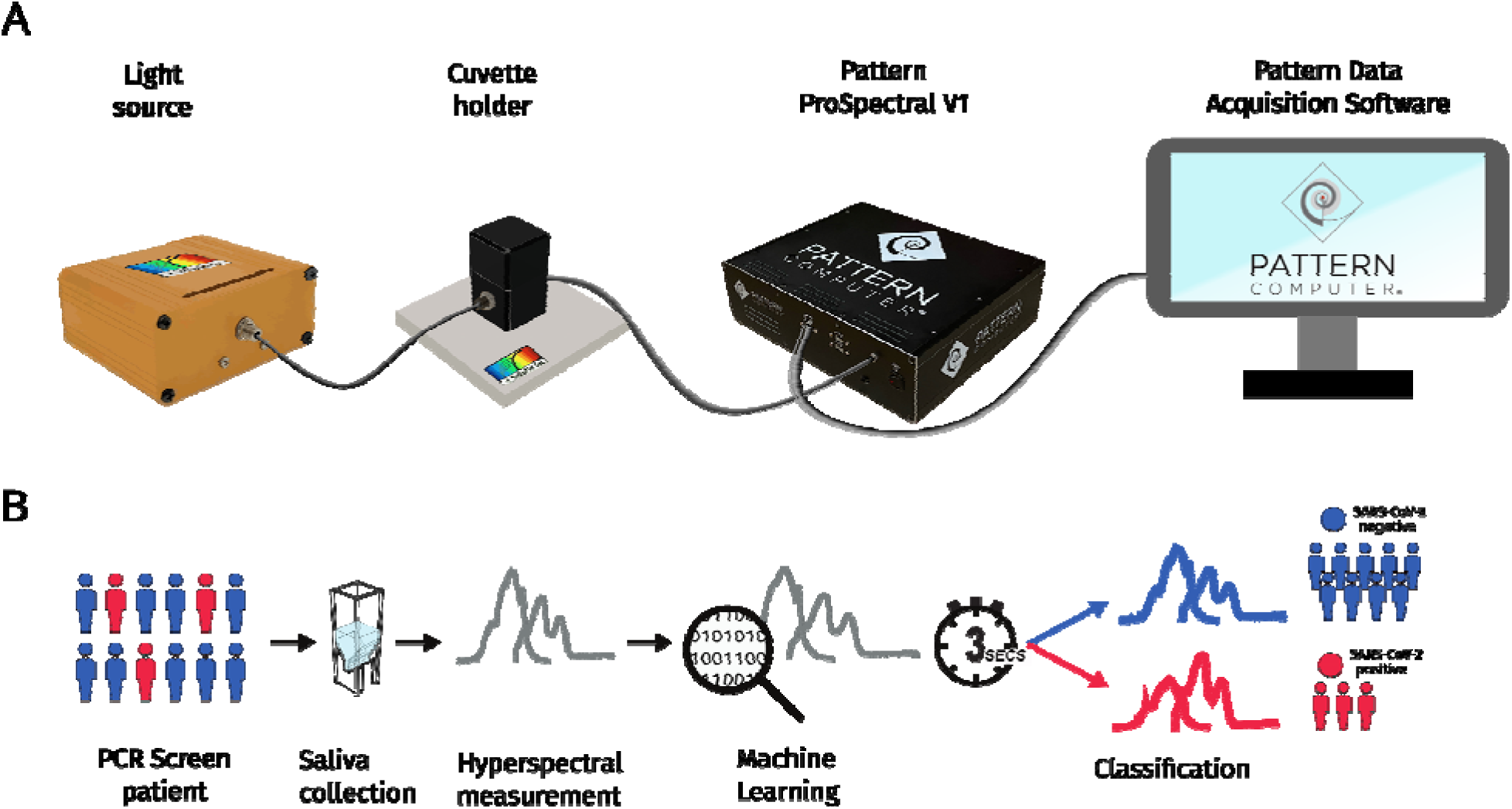
Rapid hyperspectral sensing for COVID-19 detection. A) The ProSpectral™ V1 assembled platform consists of a light source that shines light through a cuvette holder enclosing the cuvette with the saliva sample. The light is passed via fiber optic cables to the ProSpectral™ V1 platform. Custom software drivers developed by Pattern Computer Inc. enable the platform to initiate the platform and convert the spectrometer output into a data file. B) The ProSpectral™ V1 platform requires no preprocessing and generates spectra across both spectrometers within three seconds, and our machine-learning analysis finds spectral features that lead to high sensitivity and specificity.

**Figure 2.**
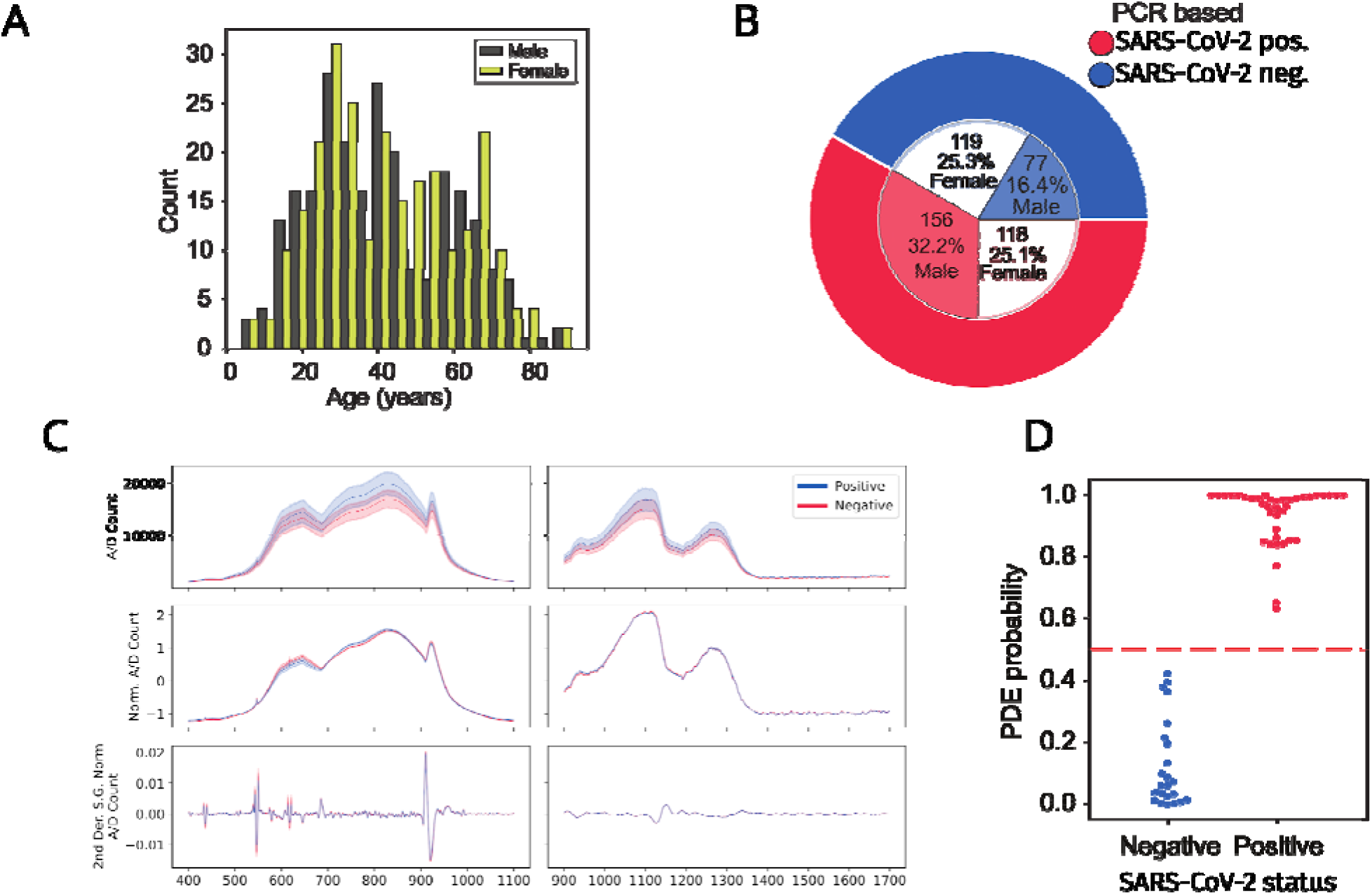
The ProSpectral™ platform can accurately distinguish between SARS-CoV-2 positive and negative saliva samples across various ages and sexes. A) The age of the frozen samples spanned from 4 to 91 sampled across both genders. B) This study’s distribution of SARS-CoV-2 positive and negative individuals is equally represented across genders. Purple represents saliva samples from male individuals, yellow represents the number of samples from female individuals. C) Overview of the spectra across both spectrometers. Spectra from SARS-CoV-2 positive saliva samples are shown in blue, and spectra from SARS-CoV-2 negative samples are shown in orange. Each line in the plot represents the average across all spectra, and the standard deviation is represented by the lighter-colored region around it. The left column contains spectra from the visible range spectrometer from 400 to 1100 nm. The right column shows data from the near-infrared spectrometer spanning 900 to 1700 nm. The y-axis represents the measurement value from the spectrometer converted to digital count. The first row is the unprocessed spectra and is the input to all the machine-learning approaches used in this study. For visualization, we also show the normalized spectra (second row) and the Savitzky-Golay 2nd derivative of the normalized spectra (window length = 21, derivative = 2, polyorder = 3) (third row). D) Across the several models we tested, the Pattern Discovery Engine^TM^ achieved perfect accuracy in classifying the holdout dataset.

To determine optimal preprocessing steps and machine learning classifiers to analyze hyperspectral measurements, and to classify patients as either SARS-CoV-2 negative or SARS-CoV-2 positive, we compared random forest, XGBoost, and H2O AutoML-based models (Supplementary Figure 1A, B). Random forest classifiers consist of an ensemble of decision trees that use randomness to create robust classification models. XGBoost is a gradient-boosting framework that uses an ensemble approach similar to random forests and has been shown to have high performance on a variety of classification tasks. Lastly, H2O AutoML is a framework that allows for training a large variety of models and includes features to evaluate these models efficiently and build ensembles of models based on the highest-performing models. We also evaluated the dataset using Pattern Computer Inc.’s suite of machine learning algorithms called the Pattern Discovery Engine™ (PDE). The PDE is a suite of proprietary algorithms developed by Pattern Computer Inc. and can discover previously unknown patterns within large datasets, without making prior assumptions, performing feature engineering, or pruning data. The PDE uses relationships within the data to build a set of directed acyclic graphs and leverage their topology to reduce dimensionality and discover hidden patterns within the data. These algorithms use the patterns they uncovered to build predictive and, when required, explainable models.

With up to ten spectral measurements for each saliva, we also wanted to explore how to aggregate the spectra or probabilities into a single likelihood of COVID-19 infection (Supplementary Figure 1C-E). For each of the mentioned models, input the data with the spectra averaged, concatentated all the measurements into a single input, or treated each of the ten as an individual sample and took the average of all the probabilities.

As noted earlier, to cover an expansive spectral range, our diagnostic platform contains two spectrometers, thereby ensuring coverage of a broad suite of biochemical targets. However, this feature also increases cost and complexity. Therefore, to quantify the benefit of incorporating two spectrometers, we compared the Receiver Operating Characteristic and Precision-Recall curves on random forest and XGBoost models trained solely on data from one of the spectrometers to those generated while using both spectrometers. (Supplementary Figure 2A-D). We found that using two spectrometers to span across the visible and near-infrared wavelengths achieved a higher classification score (Area Under the Curve (AUC)=0·97-0·98, Average Precision (AP) = 0·98) compared to using data from only one spectrometer (AUC = 0·74-0·87, AP = 0·79-0·92). Based on this, we conclude that while using two spectrometers will increase the cost and complexity, the combined data is necessary for accurate prediction of SARS-CoV-2 infection status and to make this platform competitive with the gold standard, which is PCR. Further, the tandem use of two spectrometers will ensure the ability to capture diverse biochemical signatures along the entire electromagnetic spectrum, which is required for truly agnostic sensing.

To understand the best way to convert our spectral data into biological insights, we applied four different models and multiple preprocessing approaches to determine the best overall approach to classifying COVID-19 status from saliva. Machine learning and artificial intelligence have different bias and variance trade-offs, and previous work applying models to hyperspectral sensing data shows that it’s hard to predict which one will be the best.^27^ Across all four models, we achieved high sensitivity (0·89 to 1·0) and specificity (0·74 to 1·0) across each aggregation approach (Table 1). The range of balanced accuracy metrics across all classifiers and aggregation methods was from 0·84 to 1·0. In instances where the measured spectral intensity saturated the spectrometer, a neutral density filter was used to attenuate. In the dataset where the spectra from a given sample were combined into a single feature vector, the neutral density filter resulted in 100% accuracy across all models, emphasizing the need to avoid saturating the spectrometer to realize precise outcomes. However, the H2O AutoML model did achieve 100% accuracy in samples measured with and without the neutral density filter. Lastly, PDE achieved 100% accuracy when averaging the probabilities across all spectral measurements for each of the ten replicate measurements for all samples (Figure 2D).

**Table 1.**
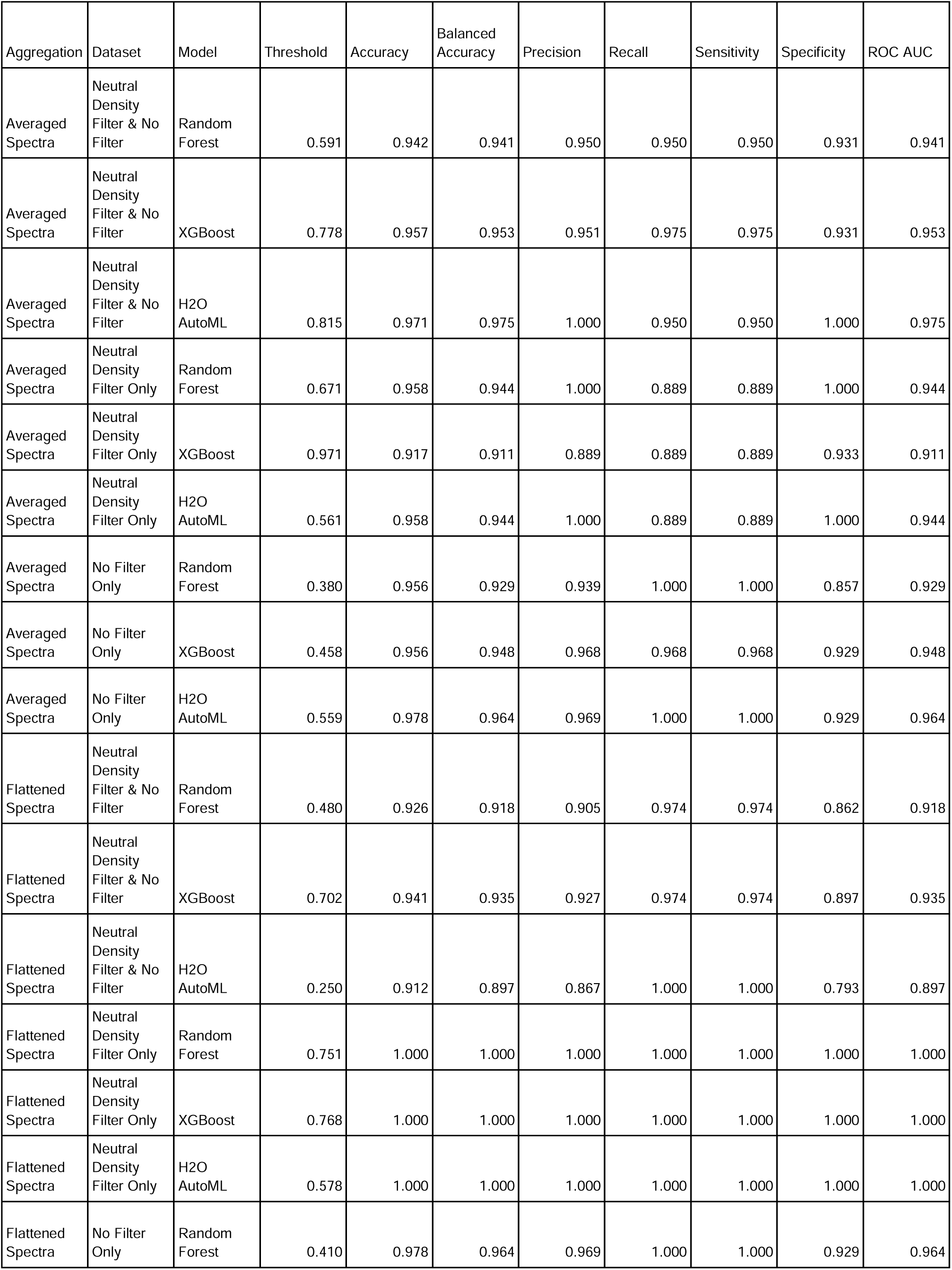
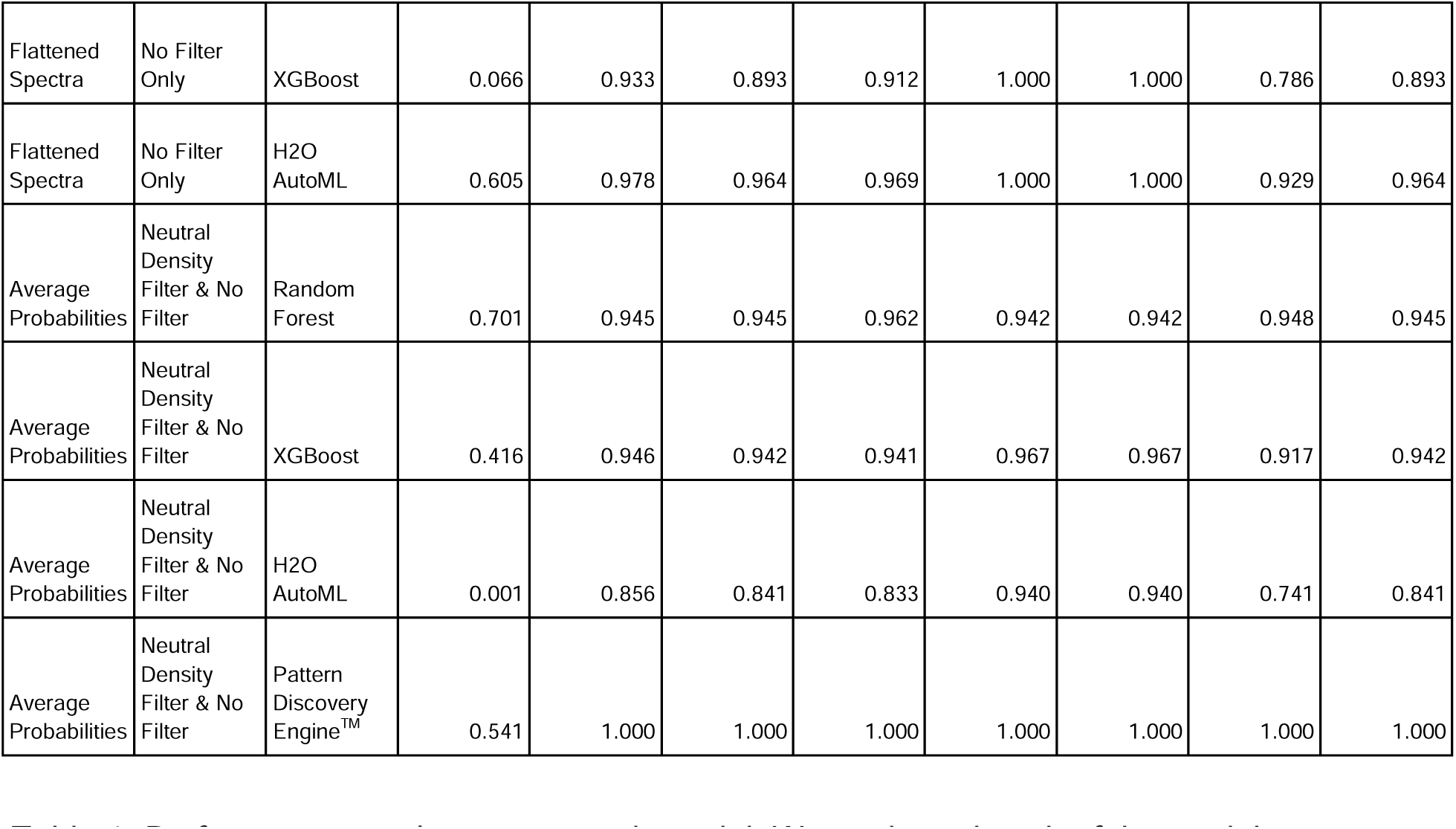
Performance metrics across each model. We evaluated each of the models across several metrics. In addition, we also evaluated the models using their default thresholds as well as the threshold that allowed no false negatives. Threshold: For the Random Forest, XGBoost, and H2O AutoML models, the maximizing F1 score threshold was used, for the PDE, it was a cutoff chosen by AUC. Data: The dataset includes samples with either only the neutral density filter, or only the no filter data, both, or both with the filter status included as an indicator variable. Aggregation: To arrive at a single probability metric for each saliva sample, we could flatten the spectra into a single array (this only includes samples where all ten spectra passed the initial 5000 filter) (flattened), average the spectra across each wavelength (averaged), or treat each single spectra of the ten spectra as a single sample, then take the average of the probabilities. Care was taken to ensure that spectra from the same sample did not leak into both the training and test datasets.

We then determined whether specific saliva-based spectral features were associated with SARS-CoV-2 infection. Our analysis identified six candidate spectral biomarkers associated with SARS-CoV-2 infection. While there was wide variation in the association of biomarkers with the models and datasets, these biomarkers were consistently observed in three model preprocessing data combinations (Supplementary Figure 2C-E). The spectral features were observed in the following wavelengths: 689 nm, 732 nm, 735 nm, 736 nm, 739 nm, and 740 nm, from the visible range spectrometer.

To generate the ground truth, we measured mRNA expression levels for SARS-CoV-2 open reading frames *ORF1a* and *ORF1b*, and the structural protein encoding *nucleocapsid* (*N*) genes, N1, and N2. For each of the saliva samples, cycle threshold (Ct) values were collected and used as the gold standard for quantifying the accuracy and sensitivity of the diagnostic prediction. The highest Ct values observed for these genes were 34·5 (*ORF1a*), 33·5 (*ORF1b*), 35·0 (*N1*), and 38·6 (*N2*), respectively, indicating low viral load for these patients. Our models could accurately classify even samples with such low viral loads, indicating high predictive power competitive with RT-PCR based methods.

Our dataset included samples from 23 individuals who were negative for SARS-CoV-2 but presented with flu-like symptoms (Supplementary Figure 3). Evaluation of these samples showed that additional spectral biomarkers were identified that were not present in the SARS-CoV-2 positive samples (Supplementary Figure 4). Although our current sample size is limited for accurate discriminative identification of other infections, these data provide preliminary support that spectral measurements have the specificity to achieve a discriminative diagnosis of related viral infections in a clinical sample.

## Discussion

We present that innovative engineering and miniaturization of hyperspectral sensors and advances in machine learning and artificial intelligence offer promising progress towards realizing reagent-free, rapid, and pathogen-agnostic diagnostics at the point of need. As the first step towards that, we demonstrate the diagnosis of COVID-19 in non-invasively collected saliva samples from 470 patients with exquisite sensitivity and specificity, irrespective of the choice of ML classifier used. Indeed, our method could identify COVID-19 even earlier than PCR-based methods in a very small subset of samples. In addition to identifying spectral signatures unique to COVID-19; we also demonstrate the feasibility of distinctive spectral patterns associated with diverse viral pathogens. These findings demonstrate the feasibility of spectral diagnostics of infection in real-time, without reagents or sample processing, offering promise, especially in global health and military applications.

As evidenced by COVID-19 and other emerging threats, there is an imminent need for rapidly developed, reagent-free, deployable diagnostics. Of the methods available, NGS is arguably the most promising agnostic approach for pathogen detection today. However, NGS requires nucleic acid extraction, library preparation, sequencing, and annotation through alignment against reference genomes, similarity searches, or assembling contiguous genomes of novel pathogens. In addition, NGS demands specialized expertise, complex sample preparation and data analysis, highly specialized equipment, and significant computational resources. The ProSpectral™ hyperspectral platform provides a complementary, rapid point-of-care device that combines reagent-free, field deployable size and a rapid (3 seconds) sensor platform with automated back-end machine-learning to immediately and accurately identify disease state. This translates to building out datasets across various pathogens and developing additional clinical applications of hyperspectral diagnostics.

The expediency of hyperspectral diagnostics is contingent on the associated machine learning and artificial intelligence algorithms that allow for sensitive signature discovery. Within highly defined diagnostic questions, such as the one described in this manuscript, it is possible to validate outcomes accurately and ensure the physiological relevance of the diagnostics. Truly realizing the potential of hyperspectral imaging for agnostic diagnostics, however, requires standardization of associated analytic pipelines. Previous work from our laboratory and others has highlighted the need to standardize data science methods when used in concert with extremely variable complex biological data.^28^ Our team is already developing experimental and *in silico* standards to benchmark clinical measurements. As noted in the manuscript, there is ambiguity in the choice of classifiers and data processing methods that can impact the outcomes of such studies. In our work, we have addressed this issue by evaluating several classifiers, and iteratively comparing outcomes; and found that these methods yielded high sensitivity and specificity, assuring us of the validity of our studies. With standardization of data analytics, and enhanced datasets of relevant metabolite signatures, hyperspectral diagnostics can change our ability to provide early and timely agnostic diagnostics, improving situational awareness and associated decision-making during emergencies.

In addition to standardized data methods, another key challenge to expanding hyperspectral diagnostics is gathering comprehensive training data. Because traditional spectroscopy approaches can take several minutes to collect data, it is time- and labor-intensive to generate large sample sizes. By having a platform that is an order of magnitude faster (>60 seconds vs. 3 seconds), as new pathogens emerge, data can be quickly gathered to build diagnostic tests efficiently. Standardization also hampers the ability to develop more comprehensive models. Previous work has explored using miniature spectrometers to develop assays, however, these works use custom hardware setups that are unstandardized and rely on software developed by the spectrometer manufacturer and not readily available.^29,30^ The platform we have presented has been developed to build custom software drivers and has been adapted into a newer version of the platform that can work with multiple types of spectrometers. With this new, commercially available platform, we are able to generate data and develop software to be readily shared and build more sophisticated models. To further help expand hyperspectral sensing to implement diagnostic tests, we are building large standardized hyperspectral libraries of various clinical matrices to address sample uncertainty and improve the feasibility of realizing such diagnostics. Further, a relatively low number of samples are needed to create/update a ProSpectral™ usable model for a new pathogen or variant, enabling faster assay development and applicability to new and emerging targets during emergencies.

To conclude, we present a deployable hyperspectral sensing platform as a potential technology to enable rapid clinical diagnosis at the point of need.

## Methods

### Saliva samples

Saliva from 240 female and 230 male individuals between the ages of 4 - 91 years (median age = 42) were purchased through Cantor BioConnect, Inc. Of the 470 samples, 196 were COVID-19 negative and 274 were COVID-19 positive, and validation was performed by PCR. Of the SARS-CoV-2 negative samples, 77 (16·5%) were male and 119 (25·5%) were female. Of the SARS-CoV-2 positive samples, 153 (32·8%) were male, and 118 (25·3%) were female. Following collection, samples were stored at -20 □.

### Hyperspectral platform hardware and setup

The ProSpectral™ V1 platform provided by Pattern Comptuer, Inc. includes multiple spectrometers, a tungsten halogen light source, and a cuvette holder with directed light passed through via multimode fiber optic cables. The range 400-1700 nm is provided by each scan performed, at 0·4 nm pixel resolution and 2·5 nm FWHM in the 400-1100 range and 1·8 nm pixel resolution and 10.0 FWHM in the range 900-1700 (IR range overlaps with VIS range).

### Hyperspectral data acquisition

Samples were thawed in a room-temperature water bath, and moved to ice before being measured immediately in batches of 20 to 40 samples. For samples received in large cups, the samples were thawed, measured, and then frozen in smaller vials. For each saliva sample, 100 µL was loaded into a cuvette (Eppendorf™ UVette™), the light was transmitted through, and spectra were generated across a 100 ms integration time. In addition, spectra were generated for several samples using a 10% neutral density filter to attenuate the signal, where the signal intensity saturates the spectrometer’s resolution. Data was corrected for this factor, as required. Reference measurements were taken with water using In total, we generated 6465 spectra measurements. The total assay time (from loading the cuvette to outcomes) for each sample was 3 seconds.

### Data Processing

For each saliva sample, data was filtered; none of the measurements across both spectrometers was greater than 5000. We also filtered wavelengths outside the manufacturer’s recommended wavelengths, reducing the visible spectrometer data to 400 - 1100 nm and 900 - 1800 nm for the infrared spectra. Data was combined in three different ways. Averaging across each wavelength, generating a single 2182 array of data points. The second was all measurements from a sample combined to generate a single array of 21820 data points. Data from the same filter was combined, and if only a subset of measurements passed the 5000 filter, we removed it from the analysis. Lastly, we treated each of the 2182 measurements individually, from ten measurements. The final probabilities were combined by taking the mean of the probabilities. For the dataset treated individually, the training test splits were generated to ensure that none of the data from a single sample leaked into both the training and test datasets. Each model was trained and tested with samples. All data processing was performed using Python 3.11 and filtering using polars version 0.20. Visualizations were performed using matplotlib (3.8.2) and seaborn (0.13.2).

### Comparison of measurements made on individual spectrometers versus combined use of both spectrometers

We performed 10-fold cross-validation using random forest and XGBoost classifiers. These models were trained on the spectra only from the visible range, only on the spectra from the near-infrared range, or trained on the combined dataset. Each classifier calculated a probability of being either SARS-CoV-2 positive or SARS-CoV-2 negative, and these were compared with the truth labels to generate precision-recall and receiver operator characteristic curves.

### Comparison of spectral biomarkers with non-COVID infections

We trained 100 random forest models on the hyperspectral data from 23 non-COVID individuals with random samples of size 23 from the SARS-CoV-2 positive or SARS-CoV-2 negative samples where the data treated each measurement as a single sample. Each set of random forests was trained to compare non-COVID infections vs SARS-CoV-2 positive, non-SARS-CoV-2 infections vs SARS-CoV-2 negative, and SARS-CoV-2 positive vs SARS-CoV-2 negative. We extracted and averaged the feature importance for each wavelength from each random forest, and took the top 25 wavelengths sorted by the averaged feature importance.

### Hyperspectral classifiers for COVID-19

We trained classifiers using Random Forest (sci-kit learn v1.4.0), XGBoost (v.2.0.3) classifier, and H2O AutoML (v3.44.0.3). The default parameters were used for the random forest and XGBoost models. For H2O AutoML, the model training was done for twenty minutes, and the best model was used for subsequent analysis. We also applied the PDE software suite to train an additional binary classifier. For each dataset, model probabilities were calculated, and metrics were calculated by using the threshold that maximized the F1 score.

## Supporting information

MetricsTable

## Data Availability

All data produced in the present study are available upon reasonable request to the authors.

## Data Sharing

All data will be made available to others in the scientific community upon request after publication of this study. The code is available at https://github.com/bsaintjo/pattern-covid-analysis.

## Author’s contributions

BSJ: Formal Analysis; Data Curation; Software; Writing; Visualization.

AWY: Formal analysis

DJ: Methodology, Investigation, Writing - Review & Editing JI: Writing, Supervision

SG: Writing - Review and Editing

MK: Conceptualization; Data curation; CM: Writing - Review & Editing

AMS: Writing, Supervision

HM: Writing; Funding Acquisition;

JZK: Methodology; Investigation; Resources; Writing - Review & Editing; Supervision; Project administration; Funding acquisition

BB: Writing, Supervision

---

**Supplementary Figure 1.**
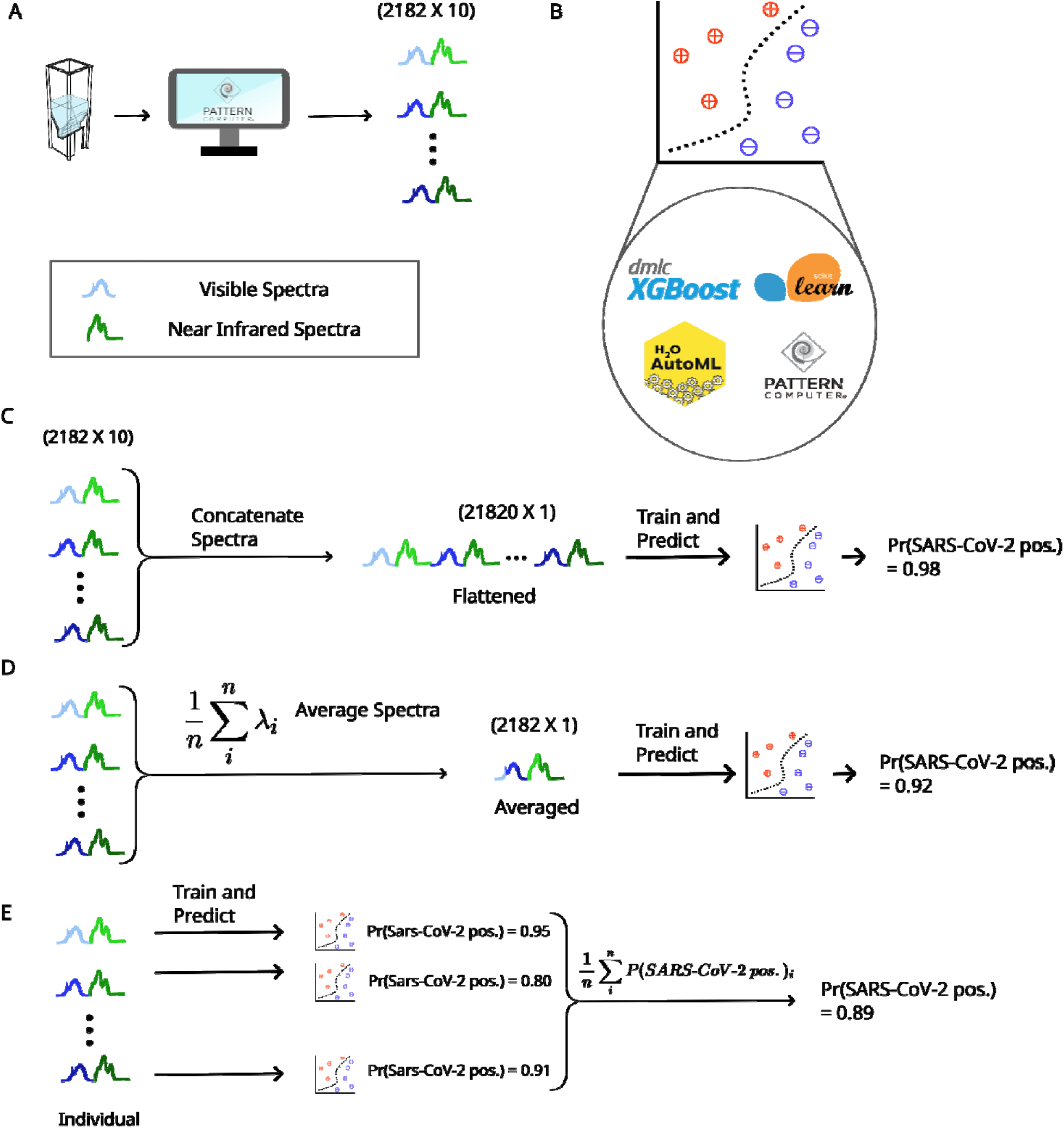
Illustration of preprocessing options. A) Overview of data generation from sample to data. The saliva sample in the cuvette is used to generate ten spectral measurements. The different hues represent one of the ten spectral measurements generated. B) We evaluated four classifiers, Random Forest from scikit-learn, XGBoost classifier, H2O AutoML classifier, and the Pattern Discovery Engine^TM^. For each of these models, they will take the input spectra(s) and output a single probability estimate. C) For flattened spectra, data from each of the ten measurements are concatenated into a single vector. This vector is then input into the models we used to produce a single probability estimate. C) For each wavelength, the spectral measurements are averaged into a single vector and input into the machine learning model. D) Each measurement is treated as an individual sample and input into the machine learning models directly. The resulting probabilities are either averaged into a single probability estimate.

**Supplementary Figure 2.**
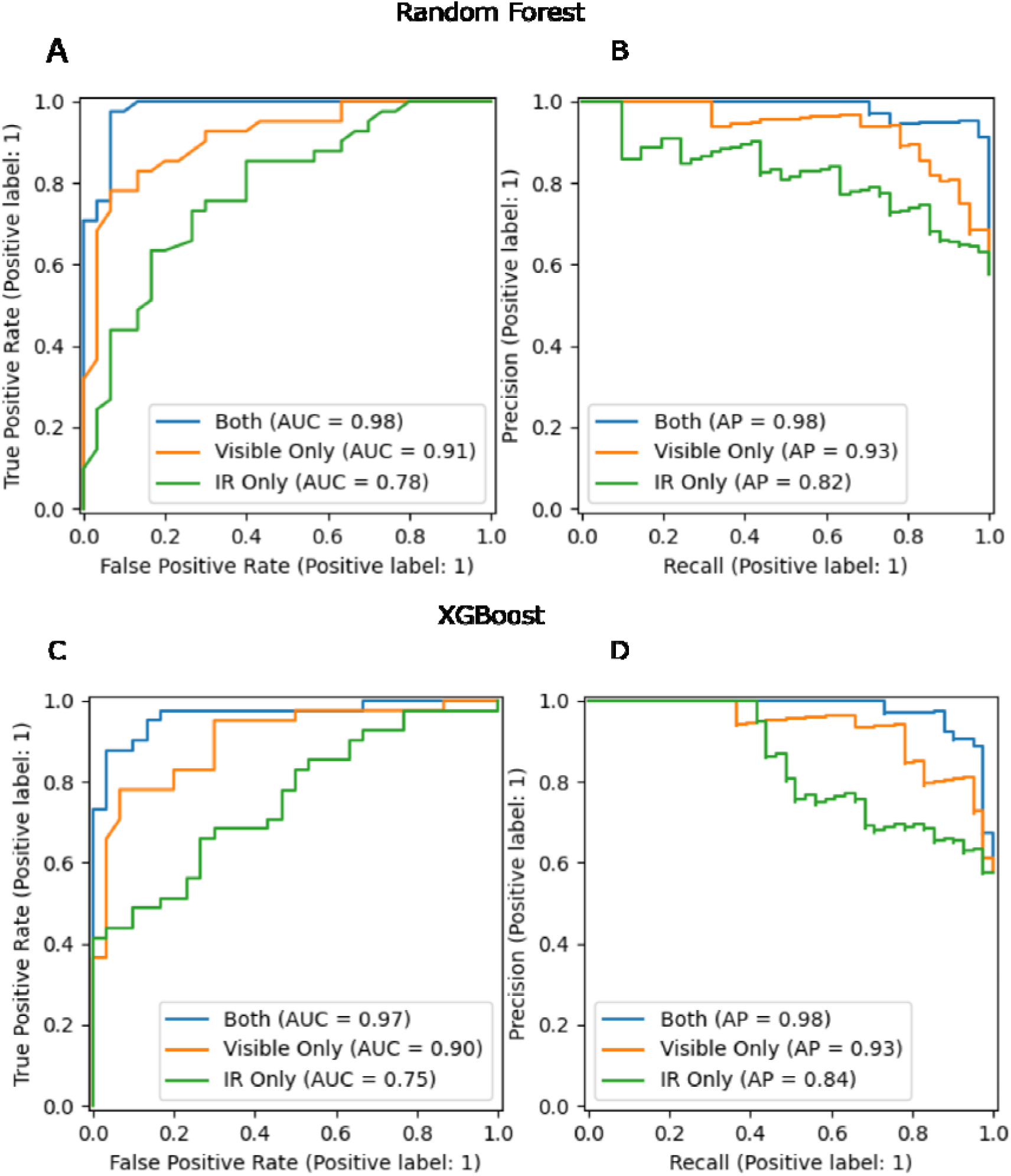
Spectra from visible and infrared are crucial for high classification performance on neutral density filter datasets. We trained a random forest classifier and XGBoost classifier model on data from only the visible, near-infrared, or both visible and near-infrared, and on the same training and test splits in the previous analyses. A) ROC and B) Precision-Recall Curves for the random forest classifier and C) ROC and D) curves for the XGBoost classifier. We found that classification is highest for either model when using spectra from both spectrometers instead of a single spectrometer. Combining both spectrometers into a single diagnostic platform is crucial for high diagnostic performance.

**Supplementary Figure 3.**
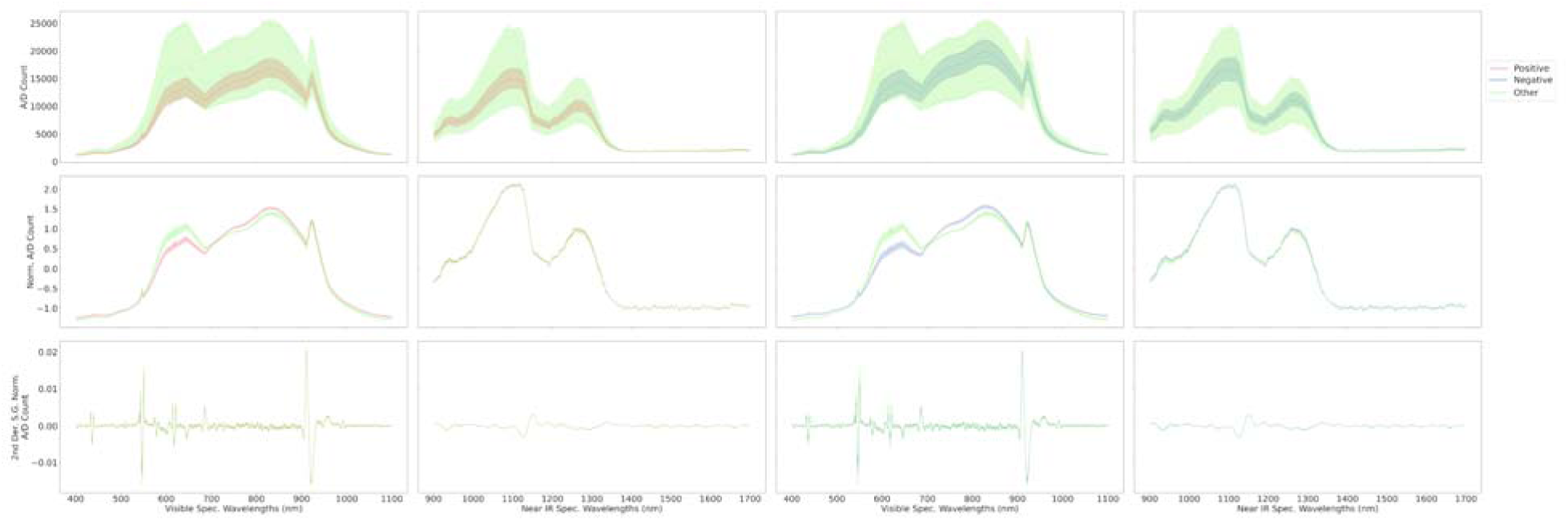
Comparison of non-SARS-CoV-2 saliva spectra with positive and negative samples shows differences across multiple wavelengths. Comparison of the SARS-CoV-2 positive spectra (red) with the spectra of saliva samples confirmed to not have SARS-CoV-2 but have some flu-like symptoms (green) is shown in the left two columns, and the comparison of the SARS-CoV-2 negative spectra (blue) with the non-SARS-CoV-2 spectra is shown in the right two columns. The first row is the unprocessed spectra and is the input to all the machine-learning approaches used in this study. For visualization, we also show the normalized spectra (second row) and the Savitzky-Golay 2nd derivative of the normalized spectra (window length = 21, derivative = 2, polyorder = 3) (third row). Overall, because of the few non-SARS-CoV-2 samples (23), there is a larger variation in the spectra and overall is different compared to both SARS-CoV-2 positive and SARS-CoV-2 negative samples.

**Supplementary Figure 4.**
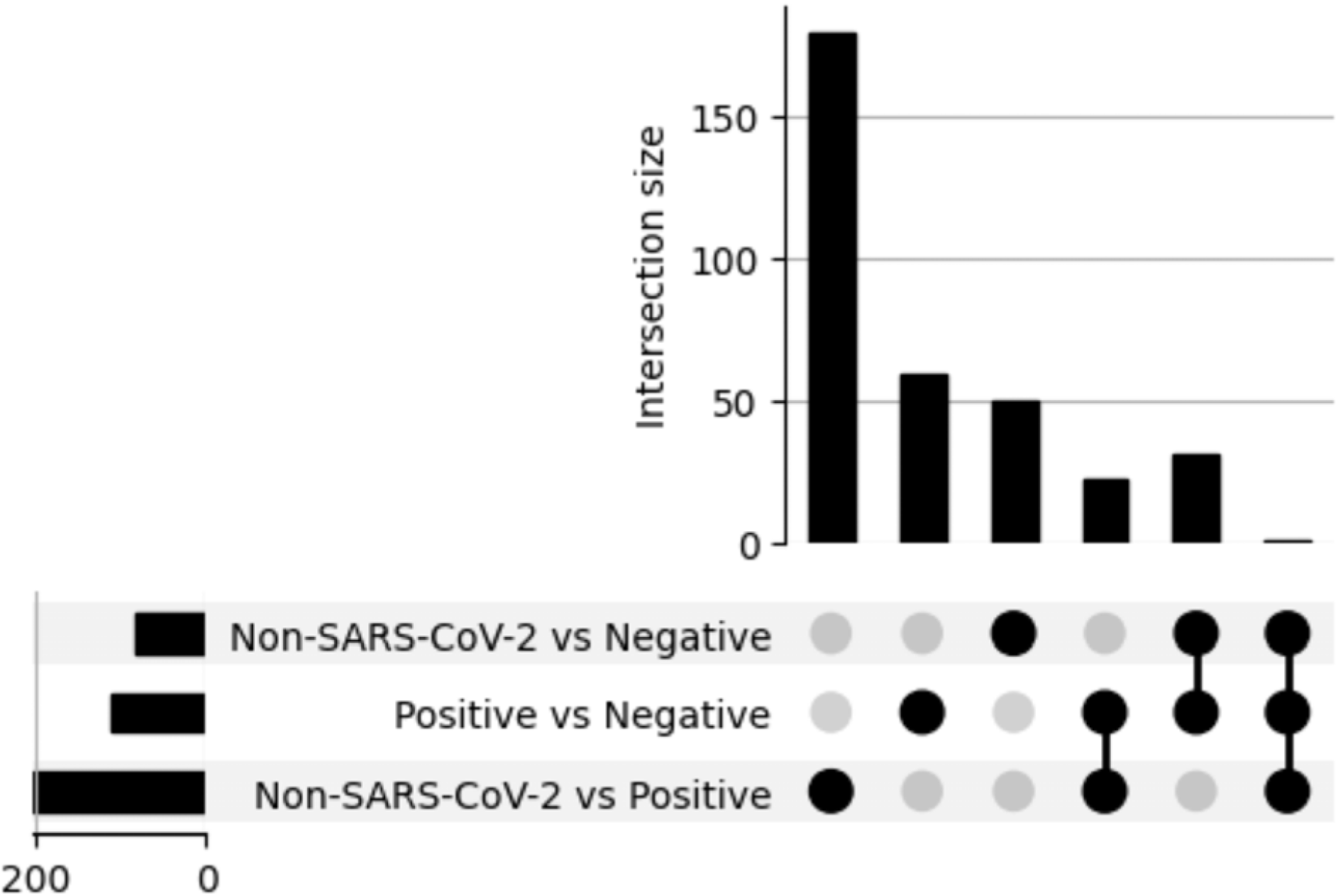
Random Forest detects spectral features that are specific to non-COVID Infections. Three sets of models were trained. Each model, the SARS-CoV-2 negative or SARS-CoV-2 positive dataset was downsampled to 23 samples. We trained 100 random forest models, comparing non-SARS-CoV-2 infections vs SARS-CoV-2 negative (row 1), SARS-CoV-2 positive vs SARS-CoV-2 negative (row 2), and non-SARS-CoV-2 infections vs SARS-CoV-2 positive (row 3). We averaged the feature importance across all 100 models and took the top 25 wavelengths ranked by the average feature importance. For these intersection plots, we also considered the wavelengths within 5 nm of the important wavelengths also as important. Each dot represents the intersection of important wavelengths for each row. Multiple dots connected by a line indicate those models had important wavelengths shared between all of them. We found that of the 25 spectral features in the non-SARS-CoV-2 infections versus SARS-CoV-2 positive model (column 1), most were specific to this model compared to non-SARS-CoV-2 infections versus SARS-CoV-2 negative (column 2) or SARS-CoV-2 positive vs SARS-CoV-2 negative (column 3). Overall, this indicates that the ProSpectral™ V1 diagnostic platform can detect spectral biomarkers that differentiate between SARS-CoV-2 positive and non-SARS-CoV-2 infections, and suggests that with more samples, we could accurately distinguish between types of infections.

## Bibliography

1 Manore C, Graham T, Carr A, et al. Modeling and Cost Benefit Analysis to Guide Deployment of POC Diagnostics for Non-typhoidal Salmonella Infections with Antimicrobial Resistance. Sci Rep 2019; 9: 11245.

2 Iliescu FS, Ionescu AM, Gogianu L, et al. Point-of-Care Testing-The Key in the Battle against SARS-CoV-2 Pandemic. Micromachines (Basel) 2021; 12. DOI:10.3390/mi12121464.

3 Kosack CS, Page A-L, Klatser PR. A guide to aid the selection of diagnostic tests. Bull World Health Organ 2017; 95: 639–45.

4 Bartlow AW, Stromberg ZR, Gleasner CD, et al. Comparing variability in diagnosis of upper respiratory tract infections in patients using syndromic, next generation sequencing, and PCR-based methods. PLOS Glob Public Health 2022; 2: e0000811.

5 Nayar G, Seabolt EE, Kunitomi M, et al. Analysis and forecasting of global real time RT-PCR primers and probes for SARS-CoV-2. Sci Rep 2021; 11: 8988.

6 Maia R, Carvalho V, Faria B, et al. Diagnosis Methods for COVID-19: A Systematic Review. Micromachines (Basel) 2022; 13. DOI:10.3390/mi13081349.

7 Mukundan H, Kumar S, Price DN, et al. Rapid detection of Mycobacterium tuberculosis biomarkers in a sandwich immunoassay format using a waveguide-based optical biosensor. Tuberculosis (Edinb) 2012; 92: 407–16.

8 Mukundan H, Kubicek JZ, Holt A, et al. Planar optical waveguide-based biosensor for the quantitative detection of tumor markers. Sensors and Actuators B: Chemical 2009; 138: 453–60.

9 Morton L, Creppage K, Rahman N, et al. Challenges and opportunities in pathogen agnostic sequencing for public health surveillance: lessons learned from the global emerging infections surveillance program. Health Secur 2024; 22: 16–24.

10 Gauthier NPG, Chorlton SD, Krajden M, Manges AR. Agnostic sequencing for detection of viral pathogens. Clin Microbiol Rev 2023; 36: e0011922.

11 Wang C, Liu M, Wang Z, Li S, Deng Y, He N. Point-of-care diagnostics for infectious diseases: From methods to devices. Nano Today 2021; 37: 101092.

12 Fani M, Zandi M, Soltani S, Abbasi S. Future developments in biosensors for field-ready SARS-CoV-2 virus diagnostics. Biotechnol Appl Biochem 2021; 68: 695–9.

13 Kitane DL, Loukman S, Marchoudi N, et al. A simple and fast spectroscopy-based technique for Covid-19 diagnosis. Sci Rep 2021; 11: 16740.

14 Barauna VG, Singh MN, Barbosa LL, et al. Ultrarapid On-Site Detection of SARS-CoV-2 Infection Using Simple ATR-FTIR Spectroscopy and an Analysis Algorithm: High Sensitivity and Specificity. Anal Chem 2021; 93: 2950–8.

15 Ahmad MA, Olule LJA, Meetani M, et al. Detection of SARS-CoV-2 in COVID-19 Patient Nasal Swab Samples Using Signal Processing. IEEE J Sel Top Signal Process 2022; 16: 164–74.

16 Nogueira MS, Leal LB, Marcarini WD, et al. Rapid diagnosis of COVID-19 using FT-IR ATR spectroscopy and machine learning. Sci Rep 2021; 11: 15409.

17 Nascimento MHC, Marcarini WD, Folli GS, et al. Noninvasive Diagnostic for COVID-19 from Saliva Biofluid via FTIR Spectroscopy and Multivariate Analysis. Anal Chem 2022; 94: 2425–33.

18 Bunaciu AA, Aboul-Enein HY. Determination of COVID-19 viruses in saliva using Fourier transform infrared spectroscopy. Chinese Journal of Analytical Chemistry 2022; 50: 100178.

19 Heino H, Rieppo L, Männistö T, Sillanpää MJ, Mäntynen V, Saarakkala S. Diagnostic performance of attenuated total reflection Fourier-transform infrared spectroscopy for detecting COVID-19 from routine nasopharyngeal swab samples. Sci Rep 2022; 12: 20358.

20 Martinez-Cuazitl A, Vazquez-Zapien GJ, Sanchez-Brito M, et al. ATR-FTIR spectrum analysis of saliva samples from COVID-19 positive patients. Sci Rep 2021; 11: 19980.

21 Kazmer ST, Hartel G, Robinson H, et al. Pathophysiological Response to SARS-CoV-2 Infection Detected by Infrared Spectroscopy Enables Rapid and Robust Saliva Screening for COVID-19. Biomedicines 2022; 10. DOI:10.3390/biomedicines10020351.

22 Lovergne L, Ghosh D, Schuck R, et al. An infrared spectral biomarker accurately predicts neurodegenerative disease class in the absence of overt symptoms. Sci Rep 2021; 11: 15598.

23 Zhu C, Fu X, Zhang J, Qin K, Wu C. Review of portable near infrared spectrometers: Current status and new techniques. J Near Infrared Spectrosc 2022; 30: 51–66.

24 Zhang C-Z, Cheng X-Q, Li J-Y, et al. Saliva in the diagnosis of diseases. Int J Oral Sci 2016; 8: 133–7.

25 Wang X, Kaczor-Urbanowicz KE, Wong DTW. Salivary biomarkers in cancer detection. Med Oncol 2017; 34: 7.

26 Arora A, Kaur D, Patiyal S, Kaur D, Tomer R, Raghava GPS. SalivaDB-a comprehensive database for salivary biomarkers in humans. Database (Oxford) 2023; 2023. DOI:10.1093/database/baad002.

27 Inman JL, Wu Y, Chen L, et al. Long-term, non-invasive FTIR detection of low-dose ionizing radiation exposure. Sci Rep 2024; 14: 6119.

28 Martinez KM, Wilding K, Llewellyn TR, et al. Factors influencing accuracy, interpretability and reproducibility in the use of machine learning in biology. Res Sq 2024; published online March 27. DOI:10.21203/rs.3.rs-4171489/v1.

29 Poh AH, Mahamd Adikan FR, Moghavvemi M, et al. Precursors to non-invasive clinical dengue screening: Multivariate signature analysis of in-vivo diffuse skin reflectance spectroscopy on febrile patients in Malaysia. PLoS ONE 2020; 15: e0228923.

30 Coelho BFO, Nunes SLP, de França CA, et al. On the feasibility of Vis-NIR spectroscopy and machine learning for real time SARS-CoV-2 detection. Spectrochim Acta A Mol Biomol Spectrosc 2024; 308: 123735.

