## SupplementaryMaterials for "Reagent-free Hyperspectral Diagnosis of SARS-CoV-2 Infection in saliva samples"

Supplementary Materials

**Supplementary Figure 1. Illustration of preprocessing options.** **2**
**Supplementary Figure 2. Spectra from visible and infrared are crucial for high classification performance on neutral density filter datasets.** **4**
**Supplementary Figure 3. Comparison of non-SARS-CoV-2 saliva spectra with positive and negative samples shows differences across multiple wavelengths.** **6**
**Supplementary Figure 4. Random Forest detects spectral features that are specific to non-COVID Infections.** **7**

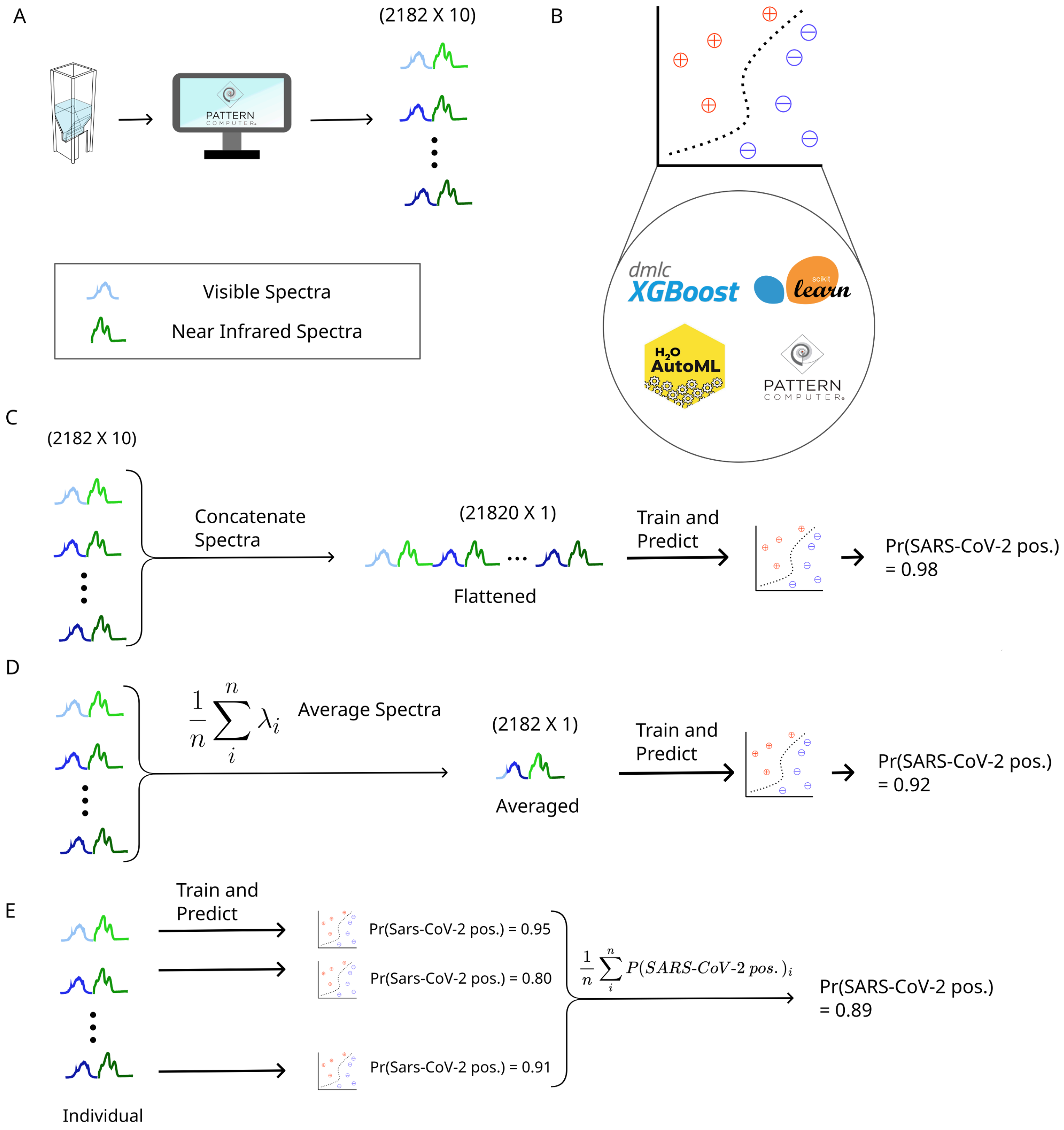


Supplementary Figure 1. Illustration of preprocessing options.

A) Overview of data generation from sample to data. The saliva sample in the cuvette is used to generate ten spectral measurements. The different hues represent one of the ten spectral measurements generated. B) We evaluated four classifiers, Random Forest from scikit-learn, XGBoost classifier, H2O AutoML classifier, and the Pattern Discovery Engine™. For each of these models, they will take the input spectra(s) and output a single probability estimate. C) For flattened spectra, data from each of the ten measurements are concatenated into a single vector. This vector is then input into the models we used to produce a single probability estimate. C) For each wavelength, the spectral measurements are averaged into a single vector and input into the machine learning model. D) Each measurement is treated as an individual sample and input into the machine learning models directly. The resulting probabilities are either averaged into a single probability estimate.

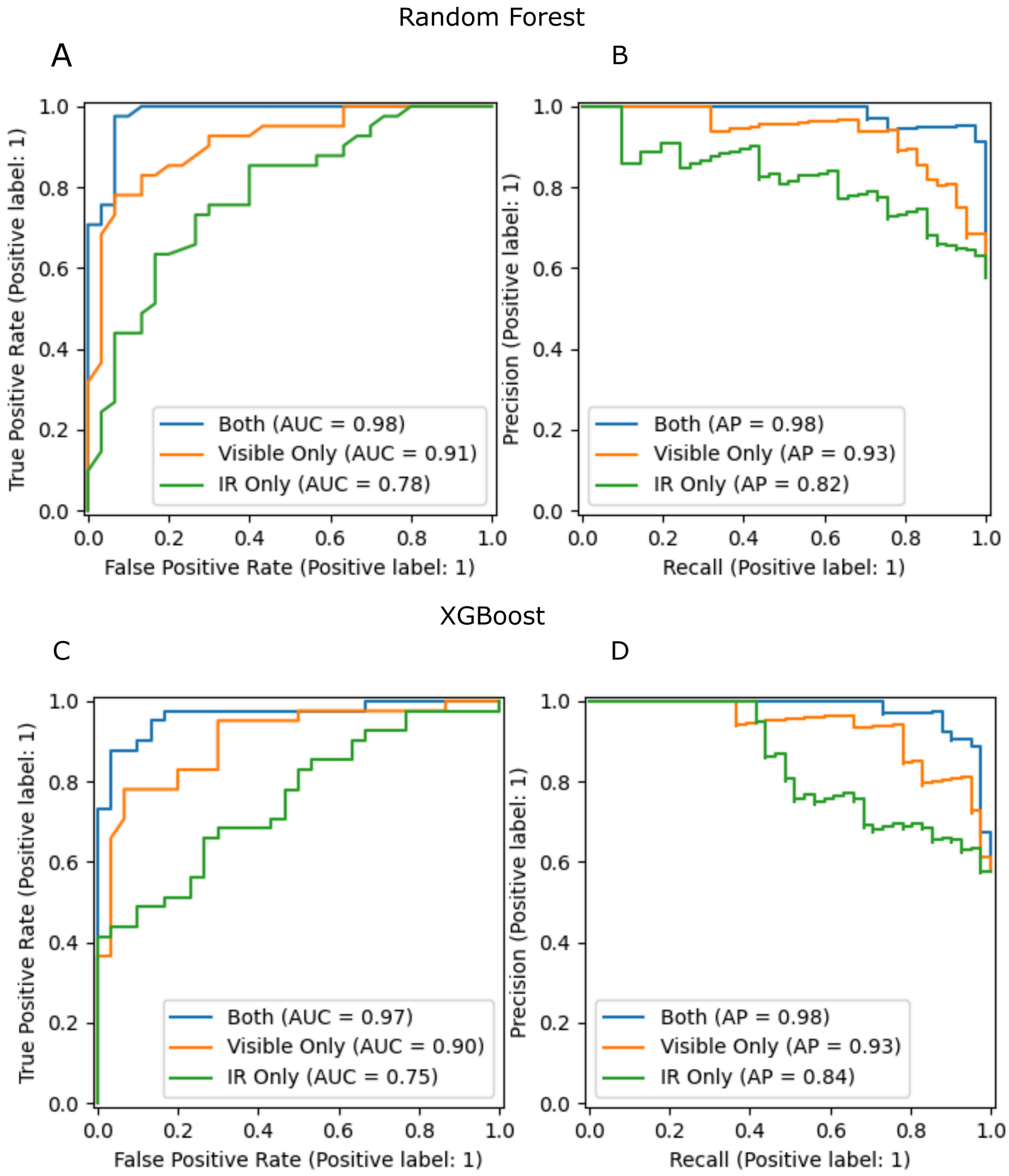


Supplementary Figure 2. Spectra from visible and infrared are crucial for high classification performance on neutral density filter datasets.

We trained a random forest classifier and XGBoost classifier model on data from only the visible, near-infrared, or both visible and near-infrared, and on the same training and test splits in the previous analyses. A) ROC and B) Precision-Recall Curves for the random forest classifier and C) ROC and D) curves for the XGBoost classifier. We found that classification is highest for either model when using spectra from both spectrometers instead of a single spectrometer. Combining both spectrometers into a single diagnostic platform is crucial for high diagnostic performance.

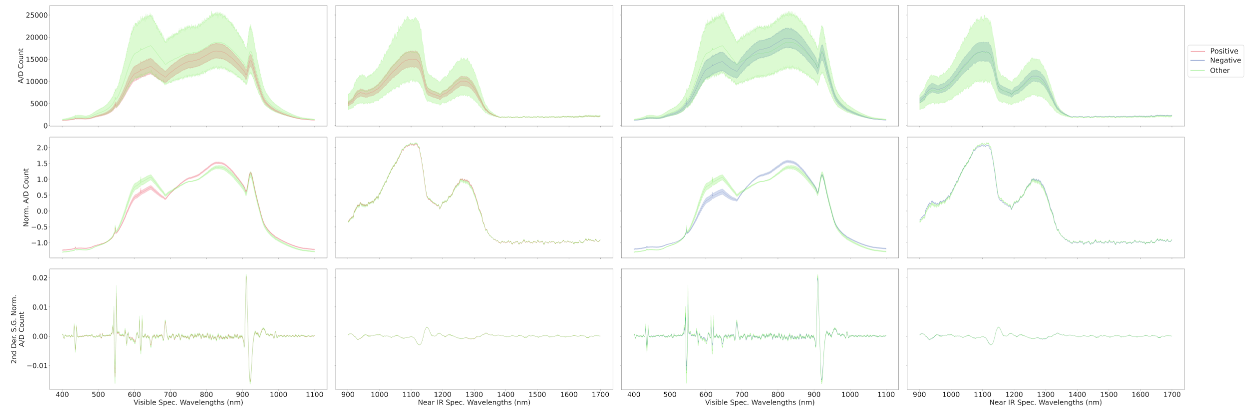


Supplementary Figure 3. Comparison of non-SARS-CoV-2 saliva spectra with positive and negative samples shows differences across multiple wavelengths.

Comparison of the SARS-CoV-2 positive spectra (red) with the spectra of saliva samples confirmed to not have SARS-CoV-2 but have some flu-like symptoms (green) is shown in the left two columns, and the comparison of the SARS-CoV-2 negative spectra (blue) with the non-SARS-CoV-2 spectra is shown in the right two columns. The first row is the unprocessed spectra and is the input to all the machine-learning approaches used in this study. For visualization, we also show the normalized spectra (second row) and the Savitzky-Golay 2nd derivative of the normalized spectra (window length = 21, derivative = 2, polyorder = 3) (third row). Overall, because of the few non-SARS-CoV-2 samples (23), there is a larger variation in the spectra and overall is different compared to both SARS-CoV-2 positive and SARS-CoV-2 negative samples.

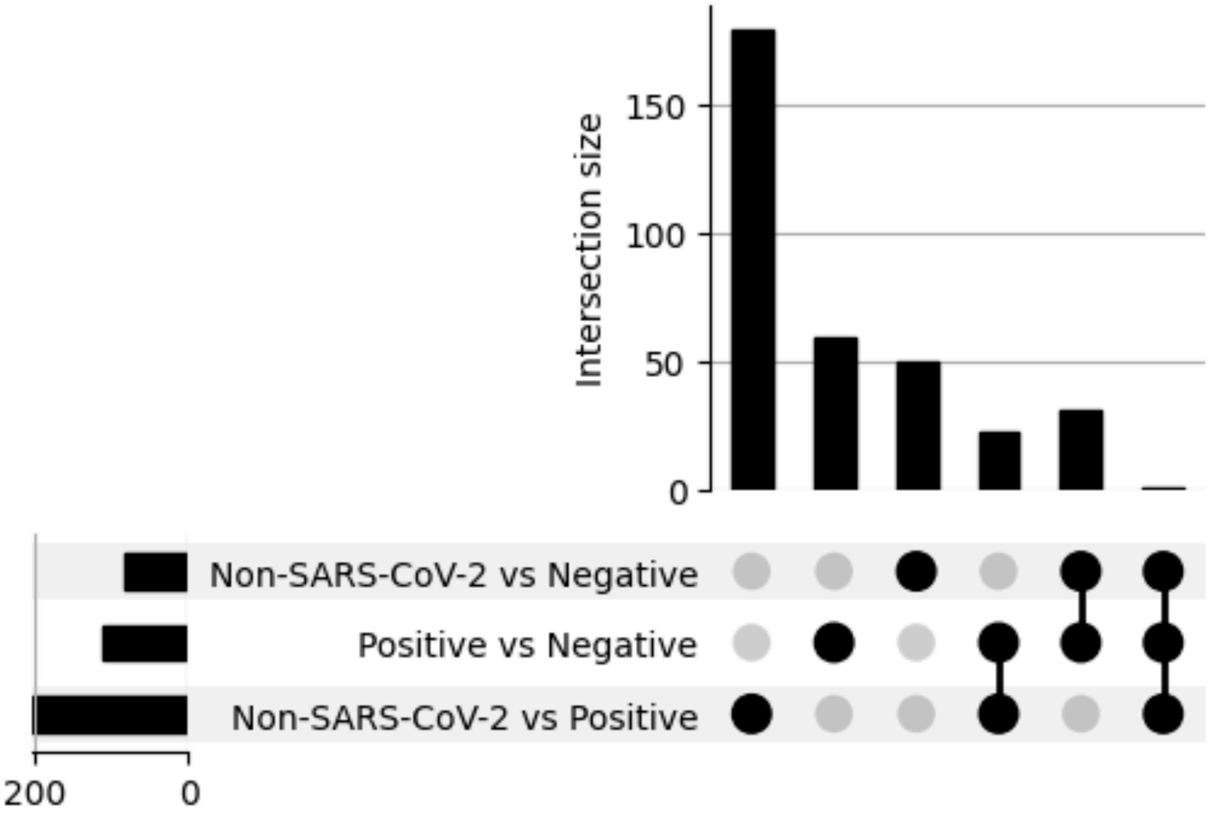


Supplementary Figure 4. Random Forest detects spectral features that are specific to non-COVID Infections.

Three sets of models were trained. Each model, the SARS-CoV-2 negative or SARS-CoV-2 positive dataset was downsampled to 23 samples. We trained 100 random forest models, comparing non-SARS-CoV-2 infections vs SARS-CoV-2 negative (row 1), SARS-CoV-2 positive vs SARS-CoV-2 negative (row 2), and non-SARS-CoV-2 infections vs SARS-CoV-2 positive (row 3). We averaged the feature importance across all 100 models and took the top 25 wavelengths ranked by the average feature importance. For these intersection plots, we also considered the wavelengths within 5 nm of the important wavelengths also as important. Each dot represents the intersection of important wavelengths for each row. Multiple dots connected by a line indicate those models had important wavelengths shared between all of them. We found that of the 25 spectral features in the non-SARS-CoV-2 infections versus SARS-CoV-2 positive model (column 1), most were specific to this model compared to non-SARS-CoV-2 infections versus SARS-CoV-2 negative (column 2) or SARS-CoV-2 positive vs SARS-CoV-2 negative (column 3). Overall, this indicates that the ProSpectral™ V1 diagnostic platform can detect spectral biomarkers that differentiate between SARS-CoV-2 positive and non-SARS-CoV-2 infections, and suggests that with more samples, we could accurately distinguish between types of infections.
