## Supplementary material for "Reagent-free Hyperspectral Diagnosis of SARS-CoV-2 Infection in saliva samples": MetricsTable

| Aggregation | Dataset | Model | Threshold | Accuracy | Balanced Accuracy | Precision | Recall | Sensitivity | Specificity | ROC AUC |
|---|---|---|---|---|---|---|---|---|---|---|
| Averaged Spectra | Neutral Density Filter & No Filter | Random Forest | 0.591 | 0.942 | 0.941 | 0.950 | 0.950 | 0.950 | 0.931 | 0.941 |
| Averaged Spectra | Neutral Density Filter & No Filter | XGBoost | 0.778 | 0.957 | 0.953 | 0.951 | 0.975 | 0.975 | 0.931 | 0.953 |
| Averaged Spectra | Neutral Density Filter & No Filter | H2O AutoML | 0.815 | 0.971 | 0.975 | 1.000 | 0.950 | 0.950 | 1.000 | 0.975 |
| Averaged Spectra | Neutral Density Filter Only | Random Forest | 0.671 | 0.958 | 0.944 | 1.000 | 0.889 | 0.889 | 1.000 | 0.944 |
| Averaged Spectra | Neutral Density Filter Only | XGBoost | 0.971 | 0.917 | 0.911 | 0.889 | 0.889 | 0.889 | 0.933 | 0.911 |
| Averaged Spectra | Neutral Density Filter Only | H2O AutoML | 0.561 | 0.958 | 0.944 | 1.000 | 0.889 | 0.889 | 1.000 | 0.944 |
| Averaged Spectra | No Filter Only | Random Forest | 0.380 | 0.956 | 0.929 | 0.939 | 1.000 | 1.000 | 0.857 | 0.929 |
| Averaged Spectra | No Filter Only | XGBoost | 0.458 | 0.956 | 0.948 | 0.968 | 0.968 | 0.968 | 0.929 | 0.948 |
| Averaged Spectra | No Filter Only | H2O AutoML | 0.559 | 0.978 | 0.964 | 0.969 | 1.000 | 1.000 | 0.929 | 0.964 |
| Flattened Spectra | Neutral Density Filter & No Filter | Random Forest | 0.480 | 0.926 | 0.918 | 0.905 | 0.974 | 0.974 | 0.862 | 0.918 |
| Flattened Spectra | Neutral Density Filter & No Filter | XGBoost | 0.702 | 0.941 | 0.935 | 0.927 | 0.974 | 0.974 | 0.897 | 0.935 |
| Flattened Spectra | Neutral Density Filter & No Filter | H2O AutoML | 0.250 | 0.912 | 0.897 | 0.867 | 1.000 | 1.000 | 0.793 | 0.897 |
| Flattened Spectra | Neutral Density Filter Only | Random Forest | 0.751 | 1.000 | 1.000 | 1.000 | 1.000 | 1.000 | 1.000 | 1.000 |
| Flattened Spectra | Neutral Density Filter Only | XGBoost | 0.768 | 1.000 | 1.000 | 1.000 | 1.000 | 1.000 | 1.000 | 1.000 |
| Flattened Spectra | Neutral Density Filter Only | H2O AutoML | 0.578 | 1.000 | 1.000 | 1.000 | 1.000 | 1.000 | 1.000 | 1.000 |
| Flattened Spectra | No Filter Only | Random Forest | 0.410 | 0.978 | 0.964 | 0.969 | 1.000 | 1.000 | 0.929 | 0.964 |
| Flattened Spectra | No Filter Only | XGBoost | 0.066 | 0.933 | 0.893 | 0.912 | 1.000 | 1.000 | 0.786 | 0.893 |

| | | | | | | | | | |
|---|---|---|---|---|---|---|---|---|---|
| Flattened Spectra | No Filter Only | H2O AutoML | 0.605 | 0.978 | 0.964 | 0.969 | 1.000 | 1.000 | 0.929 | 0.964 |
| Average Probabilities | Neutral Density Filter & No Filter | Random Forest | 0.701 | 0.945 | 0.945 | 0.962 | 0.942 | 0.942 | 0.948 | 0.945 |
| Average Probabilities | Neutral Density Filter & No Filter | XGBoost | 0.416 | 0.946 | 0.942 | 0.941 | 0.967 | 0.967 | 0.917 | 0.942 |
| Average Probabilities | Neutral Density Filter & No Filter | H2O AutoML | 0.001 | 0.856 | 0.841 | 0.833 | 0.940 | 0.940 | 0.741 | 0.841 |
| Average Probabilities | Neutral Density Filter & No Filter | Pattern Discovery Engine™ | 0.541 | 1.000 | 1.000 | 1.000 | 1.000 | 1.000 | 1.000 | 1.000 |

Table 1. Performance metrics across each model. We evaluated each of the models across several metrics. In addition, we also evaluated the models using their default thresholds as well as the threshold that allowed no false negatives. Threshold: For the Random Forest, XGBoost, and H2O AutoML models, the maximizing F1 score threshold was used, for the PDE, it was a cutoff chosen by AUC. Data: The dataset includes samples with either only the neutral density filter, or only the no filter data, both, or both with the filter status included as an indicator variable. Aggregation: To arrive at a single probability metric for each saliva sample, we could flatten the spectra into a single array (this only includes samples where all ten spectra passed the initial 5000 filter) (flattened), average the spectra across each wavelength (averaged), or treat each single spectra of the ten spectra as a single sample, then take the average of the probabilities. Care was taken to ensure that spectra from the same sample did not leak into both the training and test datasets.
